# Using wastewater sampling to investigate community-level differences in antibacterial resistance in a major urban center, USA

**DOI:** 10.1101/2024.06.21.24309301

**Authors:** Cameron Goetgeluck, Oluwatosin Olojo, Stephen P. Hilton, Orlando Sablon, Lorenzo Freeman, Patrick Person, David Clark, Robert H. Lyles, Caroline Sheikhzadeh, Marlene K. Wolfe, Maya L. Nadimpalli

## Abstract

Wastewater sampling could be a low-cost strategy for assessing patterns of antibacterial resistance across urban populations. We aimed to quantify fluoroquinolone-resistant (FQ-R) Enterobacterales, third-generation cephalosporin-resistant (3GC-R) Enterobacterales, carbapenem-resistant Enterobacterales, and the *bla*_CTX-M-15_ and *bla*_KPC_ genes in influent wastewater from 12 diverse sewersheds across metro Atlanta over 2 seasons; examine associations between culture- and digital PCR-based outcomes; and investigate relationships between sewersheds’ sociodemographic characteristics and concentrations of AR bacteria in wastewater. FQ-R Enterobacterales, 3GC-R *Escherichia coli*, and 3GC-R *Klebsiella*, *Enterobacter*, or *Citrobacter* spp. (KEC) were detected in 79-94% of samples (n=34), with concentrations differing among sewersheds. Carbapenem-resistant *E. coli* and KEC were not reliably detected. The *bla*_CTX-M-15_ gene was detected in every sample, and we detected trends towards an association with 3GC-R *E. coli* and KEC, suggesting potential utility of this gene as a monitoring target. The *bla*_KPC_ gene was detected in every sample despite carbapenem-resistant *E. coli* and KEC not reliably being detected, suggesting it is not an appropriate indicator for these bacteria. After accounting for season, repeated measures, and potential healthcare inputs, we found that sewersheds with higher proportions of crowded households, Hispanic, non-Hispanic Asian, and individuals speaking a language other than English at home had higher flow-normalized concentrations of FQ-R Enterobacterales, 3GC-R *E. coli* and/or KEC in their wastewater. Comparisons with human data are needed to determine how well sociodemographic patterns observed through wastewater mirror trends in the human population.

**Importance:** Wastewater sampling is a popular tool for the surveillance of health-related targets. Previous studies have demonstrated increases in antibiotic resistance among wastewater-derived fecal pathogens that match temporal trends in geographically-matched patient populations, indicating utility for assessing population-level patterns. Few studies have studied wastewater to examine antibiotic resistance patterns within a city, or to identify sociodemographic characteristics associated with higher concentrations of antibiotic-resistant bacteria in wastewater from certain neighborhoods. We tested municipal wastewater from 12 diverse sewersheds across metro Atlanta across two seasons. We identified significant differences in antibiotic-resistant bacterial concentrations across sewersheds, and after accounting for season, repeated sampling, and potential inputs from healthcare facilities, we found these differences were associated with community characteristics like living conditions and language. Overall, given that clinical surveillance is unlikely to be representative of the US population due to unequal healthcare access, wastewater sampling merits consideration as a novel approach for antibiotic resistance surveillance.

## Introduction

Antibacterial resistance poses a significant and growing threat to global public health. Nearly 3 million Americans suffer from antibacterial-resistant (AR) infections annually.^1^ While antibacterial resistance has been a long-standing challenge in healthcare settings, resistance among community-acquired infections is increasing, which complicates empiric treatment.

Understanding community-level patterns of antibacterial resistance is critical for guiding appropriate antibiotic treatments and monitoring antibiotic resistance trends over time. In many American cities, such patterns are difficult to assess given the disjointed network of healthcare providers, urgent care centers, hospitals, and contract laboratories that support outpatient care. Some public health departments compile these data for outpatient prescribers (e.g. NYC Department of Health and Mental Hygiene) but the vast majority do not; in these cases, outpatient prescribers often rely on antibiograms from local hospitals which may overestimate resistance in the community.^2^ More broadly, because free healthcare is not universal in the United States, using patient medical record data to understand community-level antibacterial resistance patterns is inherently biased towards individuals who can access affordable healthcare and individuals for whom doctors choose to order laboratory cultures. Finally, because patient medical records only capture active infections, the much larger fraction of persons who are gut-colonized with AR Enterobacterales like *Escherichia coli* and *Klebsiella pneumoniae* and who may therefore be at higher risk for AR community-acquired infections,^3,4^ are missed.

Wastewater surveillance could be an alternative strategy for assessing patterns of antibacterial resistance across urban populations. Regular monitoring of wastewater is becoming a standard tool for the surveillance of health-related targets.^5^ As an aggregate biological sample of entire communities, wastewater testing provides the opportunity to access information about health that is non-invasive, inexpensive, and unbiased by access to diagnostic testing resources. Importantly, wastewater can provide a composite biological sample of AR Enterobacteriales shed not only by infected persons but also gut-colonized individuals, thereby providing insight into community-level patterns of antibacterial resistance that cannot be captured by any existing alternative public health surveillance system in the US.

Metro Atlanta is a highly socioeconomically diverse urban center in the US and provides an ideal setting for exploring the feasibility of wastewater surveillance to identify community-level patterns in antibacterial resistance. More than 20% of Atlanta’s population lives in poverty (https://www.welfareinfo.org/poverty-rate/georgia/atlanta), which is almost double the US average in 2022,^6^ and exceptionally high rates of inequality exist among its racially and ethnically diverse population (https://www.atlantawealthbuilding.org/racial-wealth-gap). Here, we sought to quantify AR bacteria in wastewater across greater Atlanta, focusing on phenotypes that complicate urinary tract infection (UTI) treatment (*i.e.*, fluoroquinolone and third-generation cephalosporin [3GC] resistance), as UTIs are most often caused by gut-origin bacteria likely to be excreted in wastewater. We also considered phenotypes that are of high public health relevance (*i.e.*, carbapenem resistance). We compared culture- and digital PCR-based approaches to inform future surveillance studies, and investigated potential relationships with community-level sociodemographic characteristics related to income, race, living conditions, and education.

## Methods

### Study sites and sample collection

Untreated influent wastewater was collected from seven municipal wastewater treatment plants (WWTPs) serving greater Atlanta and five influent lines feeding into three of these WWTPs (**Figure 1**). We collected samples in December 2022, March 2023 and May 2023. WWTP-level samples were 24-hour, composite, flow-proportional samples, while influent line samples were grab samples collected using 500 mL or 1 L Nalgene bottles. Wastewater samples were transported to the Rollins School of Public Health on ice and stored at 4°C until processing, typically within one hour.

**Figure 1.**
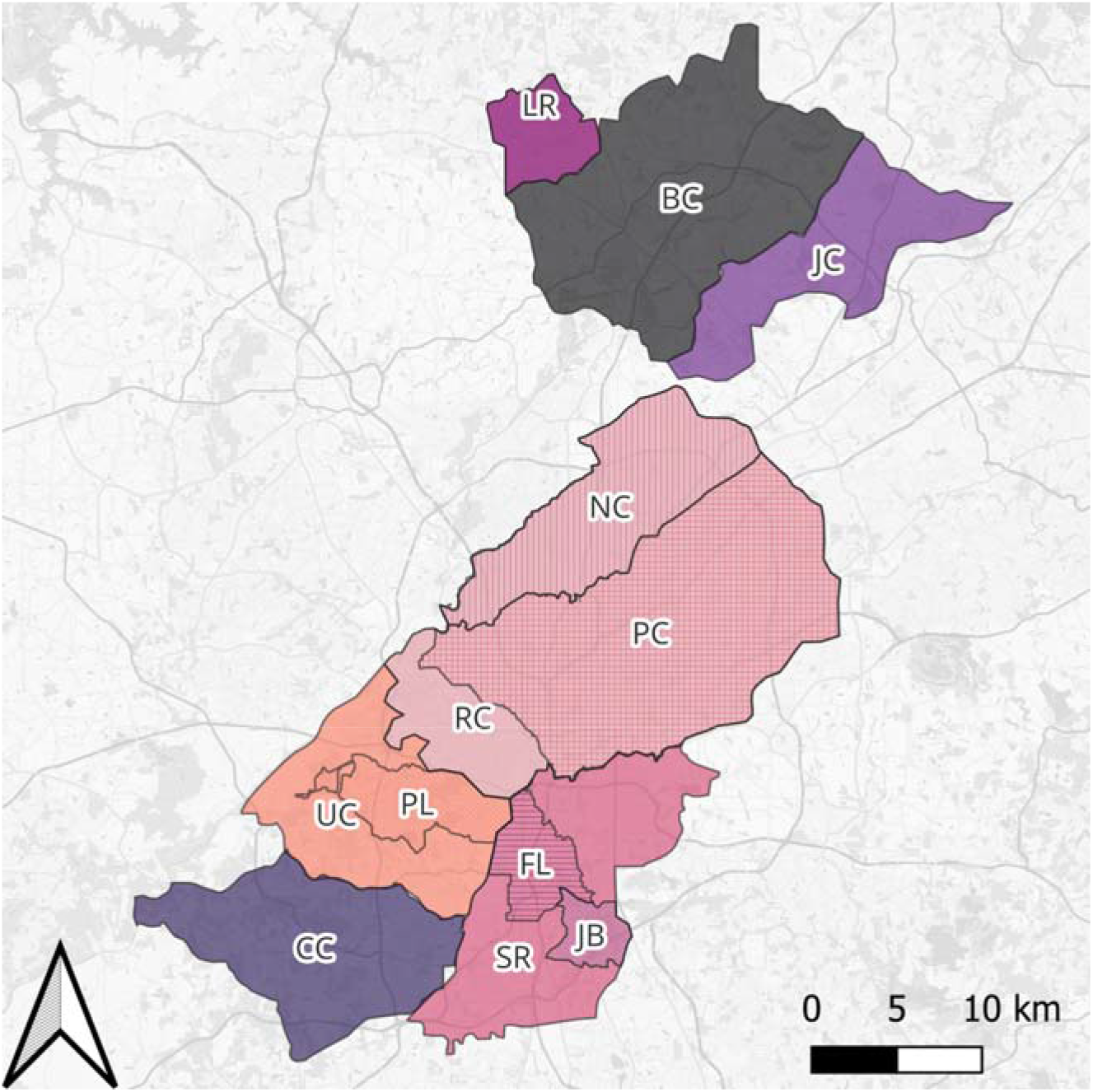
Map of sewersheds served by seven municipal wastewater treatment plants (WWTPs) and five influent lines sampled for this study. WWTPs include (starting at top, moving clockwise): LR=Little River, BC=Big Creek, JC=Johns Creek, RM=RMC Clayton, SR=South River, CC=Camp Creek, and UC=Utoy Creek. Influent lines include two influent lines of RC, NC=Nancy Creek and PC=Peachtree Creek; two influent lines of SR, FL=Flint, JB=Jonesboro, and one influent lines of UC, PL=Phillip Lee. Atlanta sewershed shapefiles were obtained from the Atlanta Department of Watershed Management, and any areas outside city boundaries were estimated by digitizing maps from an online report (Dekalb County, 2022). Shapefiles for Fulton County were obtained from the Fulton County Department of Water Services.

### Culture-based detection of antibiotic-resistant Enterobacterales

We aimed to isolate FQ-R, 3GC-R, and carbapenem-resistant Enterobacterales from wastewater using differential media. To select FQ-R Enterobacterales, we supplemented MacConkey agar with 4 µg/ml of ciprofloxacin (FQ plates). We used CHROMagar ESBL (CHROMagar Microbiology, Paris, France) to select 3GC-R *Escherichia coli* and KEC (*Klebsiella* spp., *Enterobacter* spp., and *Citrobacter* spp.) and CHROMagar mSuperCARBA (CHROMagar Microbiology, Paris, France) to select carbapenem-resistant *E. coli* and KEC.

We screened samples received in December 2022 using membrane filtration. Briefly, wastewater was homogenized by shaking bottles laterally for approximately one minute. Fifty mL were extracted and centrifuged for 15 minutes at 4000 rpm. We removed 1 mL of supernatant and prepared 10-fold serial dilutions using phosphate-buffered saline (PBS). Each dilution was briefly vortexed before being vacuum filtered in duplicate through 0.45 *µ*m membrane filters (Fisherbrand, Fisher Scientific, USA). Membrane filters were then aseptically transferred onto FQ, mSuperCarba and CHROMagar ESBL plates. We used 1:10 through 1:1000 dilutions of untreated influent wastewater to screen for 3GC- and carbapanem-resistant Enterobacterales, and 1:100 through 1:10,000 dilutions to screen for FQ-R Enterobacterales. Plates were incubated for 18 to 24 hours at 37°C.

Given that we detected many of the outcomes of interest at high concentrations in December samples, we directly plated wastewater collected in March and May 2023. Briefly, untreated wastewater was left to settle at 4°C for approximately 45 minutes upon arrival in the lab. We prepared 1:5 and 1:10 dilutions in PBS, then plated 100 µl of each in duplicate on FQ, CHROMagar ESBL, and mSuperCarba plates. Plates were incubated for 18 to 24 hours at 37°C.

We preferentially counted plates with 25-250 colony forming units (CFUs). Both tan (lactase non-producing) and pink (lactase-producing) colonies were counted on FQ plates. Pink (*E. coli*) and blue (KEC) colonies were counted on mSuperCarba and CHROMagar ESBL. For the FQ plates, our estimated lower limit of detection using either membrane filtration or a direct plating approach was 3 log_10_ CFU/100 mL. For mSuperCarba and CHROMagar ESBL, our estimated lower level of detection was 2 log_10_ CFU/100 mL when using membrane filtration and 3 log_10_ CFU/100 mL when directly plating.

### Species confirmation of cultured isolates

Up to three colonies of each desired phenotype were isolated and archived to confirm species using endpoint PCR. To confirm *E. coli* species, pink colonies from mSuperCarba and CHROMagar ESBL and both pink and tan colonies from FQ plates were screened for the *lacZ* and *yaiO* genes;^7^ isolates that harbored both genes were considered *E. coli*. All blue colonies from mSuperCarba and CHROMagar ESBL were tested for the presence of the *zikr* gene, which is specific to *K. pneumoniae*.^8^ PCR products were visualized using gel electrophoresis with 1.5% agarose gels stained with SYBR Safe.

We noted that no pink colonies on MSuperCarba were confirmed to be *E. coli* and almost no blue colonies were confirmed to be *K. pneumoniae* (**Table S1**), the primary species of interest for this study. Given these results, findings from CHROMagar mSuperCARBA are not presented.

### Molecular detection of antibiotic resistance genes in wastewater

We used clinical surveillance data from the CDC’s Emerging Infections Program (EIP), in which Georgia participates, to inform our selection of ARG alleles to measure. EIP data indicate that 97% of 3GC-R infections in EIP catchment areas harbor an extended-spectrum beta lactamase (ESBL), most commonly encoded by *bla*_CTX-M-15_.^9^ Site data from the Georgia EIP indicate that approximately 20% of carbapenem-resistant infections in greater Atlanta harbor a carbapenemase (**Table S2**), of which *bla*_KPC_ is the most common allele.^10^ Thus, we chose to measure *bla*_CTX-M-15_ and *bla*_KPC_ concentrations in wastewater. We did not measure ARGs conferring resistant to fluoroquinolone since high-level fluoroquinolone resistance (minimum inhibitory concentration >=4 ug/L) is typically mediated through chromosomal mutations of the QRDR region, rather than ARGs.^11^

We processed 9.6 mL of each wastewater sample to screen for *bla*_CTX-M-15_, *bla*_KPC_ and 16S rRNA gene. Each sample was supplemented with 150 µL each of CERES Nanotrap® Microbiome A and B and 100 µL of CERES Nanotrap® Enhancement Reagent 3 to enhance our capacity to concentrate and capture bacteria, following established protocols (APP-075). Supplemented samples were concentrated and extracted on the KingFisher™ Apex automated purification system (Thermo Scientific) utilizing the MagMAX™ Microbiome Ultra Nucleic Acid Isolation kit (Applied Biosystems™). We prepared 1:10,000 dilutions of overnight cultures of AR-BANK#0109 and ATCC BAA-1705 to use as positive controls for the *bla*_CTX-M-15_ and *bla*_KPC_ genes, respectively; these were concentrated and extracted using the same procedures. The Nanotrap script for concentration & extraction using the KingFisher™ Apex was provided by CERES Nanosciences, Inc.

To quantify *bla*_CTX-M-15_, *bla*_KPC_, and the bacterial 16S rRNA gene in wastewater, a hydrolysis probe-based approach utilizing end-point PCR along with partitioning was performed using the Qiagen QIAcuity® digital PCR platform. We used previously published primers and probe sequences (**Table S3**) and a ready to use QIAcuity® Probe PCR kit. All reactions were run on a QIAcuity® Four system using QIAcuity® Nanoplate 26k 24-well plates with a QIAcuity® Nanoplate Seal. For *bla*_CTX-M-15_ and *bla*_KPC_, we used 10 µl 4X Probe PCR Master Mix, 5 µl of enhanced GC, 3 µl of primer/probe mix, 12 µl of RNase-Free water, and 10 µl of diluted template DNA (1:10) for a total reaction volume of 40 µl per reaction per manufacturer’s recommendations. For 16S rRNA, 10 µl 4X Probe PCR Master Mix, 5 µl of enhanced GC, 2 µl of primer/probe mix, 13 µl of RNase-Free water, and 10 µl of diluted template DNA (1:10000) were used for a total reaction volume of 40 µl per reaction. For both the *bla*_CTX-M-15_ and *bla*_KPC_ targets, final concentrations for forward and reverse primers were 600 nM while the final concentration of probe was 300 nM. For the 16S rRNA target, the final concentration for forward and reverse primers were 400 nM while the probe was 200 nM. The QIAcuity dPCR cycling parameters were based on the QIAcuity® Probe PCR kit Quick-Start Protocol with modifications made to the 2-step cycling condition to fully optimize PCR conditions for each target. The dPCR cycling parameters for *bla*_CTX-M-15_ were 95°C for 2 minutes, followed by 40 cycles at 95°C for 30 seconds and 62°C for 15 seconds. For *bla*_KPC_ & 16S rRNA, dPCR cycling parameters were 95°C for 2 minutes, followed by 40 cycles at 95°C for 15 seconds and 57°C for 30 seconds.

Analysis was conducted on the QIAcuity Software Suite (Qiagen, version 2.2.026) under 1D scatterplot for absolute quantification. Common thresholds were set based on target fluorescence intensity between background/negative partitions and positive partitions relative to the positive control.

### Characterization of the *bla*_CTX-M_ allele

Colonies originally isolated on CHROMagar ESBL that were confirmed to be either *E. coli* or *K. pneumoniae* were tested for the presence of the *bla*_CTX-M_ gene using methods previously described.^6^ We selected a random subset of up to 3 *bla*_CTX-M_-harboring *E. coli* and 3 *K. pneumoniae* per sewershed, per round, for sequencing. We sequenced the forward strand of the *bla*_CTX-M_ amplicon using Sanger sequencing and determined whether a *bla*_CTX-M_ allele belonging to group 1, 2, 8, 9, 25, 64, 151, or 137 was harbored using BLASTn. We used a 98% identity threshold to identify matches.

### Demarcation of sewersheds

Sewershed shapefiles for Atlanta WWTPs (RMC Clayton [RC], South River [SR], and Utoy Creek [UC]) and two influent lines (Peachtreek Creek [PC] and Nancy Creek [NC], both of RC) were obtained from the Atlanta Department of Watershed Management, and shapefiles for Fulton County WWTPs (Little River [LR], Big Creek [BC], Johns Creek [JC}, and Camp Creek [CC]) were obtained from the Fulton County Department of Water Resources. Shapefiles for Atlanta WWTP sewersheds only provided information within the city of Atlanta. The area covered by sewersheds that extended outside the city of Atlanta was estimated using manhole data from Fulton County and by digitizing maps from a Dekalb County report.^12^

For influent lines for which shapefiles were not available (i.e., Flint [FL], Jonesboro [JB], and Phillip Lee [PL]), sewersheds were initially estimated using a network approach that identifies all manholes upstream of a collection point.^13^ This only estimated coverage within the city of Atlanta, which was refined using shape files from the Atlanta Department of Watershed Management, when available. Areas outside Atlanta were estimated by digitizing a map from a Dekalb County report.^12^

### Identification of hospitals and LTCFs within sewersheds

We identified active hospitals (https://data.cms.gov/provider-data/dataset/xubh-q36u) and LTCFs (https://data.cms.gov/provider-data/dataset/4pq5-n9py) registered with the Centers for Medicare & Medicaid Services (CMS) as of 2024. We tabulated the number of staffed beds in each facility using publicly available data from CMS or from the American Hospital Directory (https://www.ahd.com/). We identified facilities located within the boundaries of each sewershed using geocoded addresses. We used QGIS (www.qgis.org) to evaluate our confidence in each assignment. High confidence assignments were squarely within the boundaries of a given sewershed or connected to sewer mains associated with certain sewersheds, while low confidence assignments were relatively close to sewershed boundaries. The number of tabulated hospital and LTCF beds per sewershed is reported in **Figure S3**.

### Allocating census data to sewersheds

We used block group-level data from the US Census Bureau’s 2016-2021 American Community Survey^14^ to tabulate various sewershed-level characteristics of the catchment areas served by the WWTPs and influent lines we sampled. ACS data were downloaded using the TidyCensus package in R.^15^ Data from block groups allocated to a sewershed were aggregated to create summary statistics for each sewershed. When a block group was only partially within a sewershed, census variables were allocated to the sewershed using an area and population interpolation approach similar to Logan et al. 2014.^16^ Briefly, the allocation ratio for the block group was estimated using the populations from blocks, which are smaller than block groups.^17^ When a block itself was only partially within a sewershed, census variables were allocated using the ratio of the area within the sewershed. This approach assumes that sociodemographic variables measured by the US Census are homogenous within a block.

The following characteristics were aggregated for each sewershed (**Table S4):** total population; median household income in the past 12 months (in 2021 inflation-adjusted dollars); percentage of population 5 years and over who speaks languages other than English at home; percentage of population having Medicaid/means-tested public coverage only; percentage of population who is uninsured; percentage of population over 18 for whom poverty status is determined and whose income in the past 12 months was below poverty level; percentage of residents over 25 who have completed high school or an equivalent degree; percentage of population living in crowded conditions (i.e., >1.01 persons/room, not including bathrooms and kitchens); percentage of population who are Hispanic or Latino; percentage of population who are non-Hispanic White alone; percentage of population who are non-Hispanic Black alone; and percentage of population who are non-Hispanic Asian alone.

### Statistical Analysis

Descriptive statistics were compiled to examine concentrations of culture-based outcomes and ARGs, overall and per round of sampling. We normalized concentrations of AR Enterobacterales, *bla*_CTX-M-15_, and *bla*_KPC_ detected in municipal WWTP influent by daily flow rates for each sampling date, as reported by City of Atlanta and Fulton County plant operators (**Table S5**). Daily flow rates approximate the total population contributing to a 24-hour composite sample. Due to limitations in data availability, outcomes measured in influent line grab samples were normalized by average flow rates across the three months of sampling. We additionally normalized ARG concentrations by concentrations of the bacterial 16S rRNA gene.

We examined differences in the mean flow-normalized concentrations of each culture-based outcome (*i.e.*, FQ-R Enterobacterales, 3GC-R *E. coli*, 3GC-RKEC) and each ARG (*i.e.*, *bla*_CTX-M-15_, and *bla*_KPC_) between WWTP sewersheds and between rounds using a two-way analysis of variance (ANOVA) and post-hoc Tukey tests. This modeling approach treats observations as independent after conditioning on the sewershed.

We examined whether concentrations of culture-based outcomes and their corresponding ARGs (i.e., 3GC-R *E. coli* or KEC with *bla*_CTX-M-15_) were associated using a three-step modeling approach, accounting for repeated measures within each sewershed. Separate regressions were fit with 3GC-R *E. coli* and 3GC-R KEC as the outcome variables, treating *bla*_CTX-M-15_ as the predictor of interest. Indicator variables for the sampling rounds were included based on reference cell coding, together with their interactions with *bla*_CTX-M-15_, in order to investigate differences in the regression relationship across rounds. For each of the two models, we first tested the null hypothesis that the sampling round had no effect on the mean outcome (*i.e.*, coincident regressions). If this hypothesis was rejected, we then tested the null hypothesis of parallel regressions, *i.e.*, allowing the mean outcome to vary across sampling rounds but maintaining the same level of association with *bla*_CTX-M-15_ regardless of the round. In the models used to test for coincidence and parallelism, we utilized an unstructured error covariance matrix to account for correlations due to repeated measures. Subsequently, we used Akaike’s information criterion (AIC)^18^ to select the error covariance structure for the final models, with the candidates including the unstructured, compound symmetric, independence, and diagonal but heteroscedastic structures. Based on the selected model (which exhibited parallelism in each case), we assessed the magnitude of association between the outcome and *bla*_CTX-M-15_, while also examining pairwise Bonferroni-corrected comparisons of model intercepts across rounds. As with the initial repeated measures ANOVA models looking at differences across sampling round, estimation was conducted via restricted maximum likelihood as implemented in the SAS MIXED procedure.^19^

We examined whether the number of staffed hospital beds or the number of certified LTCF beds in a sewershed were associated with differences in the flow-normalized concentrations of each culture-based outcome across sewersheds using linear regression models with a GEE and an independence correlation matrix, controlling for repeated measures within each sewershed and sampling round. We performed a sensitivity analysis that excluded low confidence facility assignments; results were similar and not presented. Both the number of staffed hospital beds and the number of certified LTCF beds in a sewershed were significantly associated with concentrations of each culture-based outcome in a sewershed’s wastewater, although effect sizes were uniformly near zero (**Table S6**). To build the most parsimonious model given our limited number of observations, we thus compiled the number of staffed hospital beds and the number of certified LTCF beds into a single variable for subsequent analyses.

Lastly, we used linear regression models with a GEE and an independence correlation matrix to evaluate associations between each sewershed-level characteristic in **Table S4** and each of our culture-based outcomes of interest, controlling for repeated measures within each sewershed, sampling round, and number of hospital or LTCF beds within each sewershed.

All analyses were conducted in RStudio (Version 2022.12.0+353) or SAS 9.4 (Cary, NC).

## Results

### AR Enterobacterales and ARGs were frequently detected in municipal wastewater

Fluoroquinolone- and 3GC-R Enterobacterales were detected in the majority of wastewater samples (**Table 1**). As noted above, given that the majority of pink and blue colonies we counted on MSuperCarba plates were not confirmed to be either *E. coli* nor *K. pneumoniae* (**Table S1**), respectively, concentrations of carbapenem-resistant *E. coli* and KEC are not reported.

**Table 1.** Detection of AR Enterobacterales in Atlanta area wastewater.

| Outcome assessed | No. of samples tested (N) | No. of positive samples (n, %) | Median log<sub>10</sub>CFU per 100 mL [minimum, maximum] | Flow-normalized median log<sub>10</sub>CFU per day [minimum, maximum] |
| --- | --- | --- | --- | --- |
| 3GC-R *E. coli* | 34 | 27 (79.4) | 4.47 [1.70, 5.82] | 15.31 [11.21, 17.20] |
| 3GC-R KEC | 34 | 29 (85.2) | 4.39 [1.70, 6.69] | 15.51 [11.21, 17.54] |
| FQ-R Enterobacterales | 34 | 32 (94.1) | 4.55 [2.70, 6.32] | 15.57 [12.21, 17.17] |

*Note:* 3GC-R=third-generation cephalosporin-resistant, FQ-R=fluoroquinolone-resistant, KEC=*Klebsiella* spp., *Enterobacter* spp., *Citrobacter* spp. Findings for carbapenem-resistant Enterobacterales are not depicted because of low confidence in results.

Flow-normalized concentrations of 3GC-R *E. coli* and FQ-R Enterobacterales significantly differed across WWTP sewersheds (p<0.05 by two-way ANOVA), when accounting for sampling round (**Figure 2**). Lower concentrations of both outcomes in Little River WWTP wastewater appeared to drive these global differences. Specifically, concentrations of 3GC-R *E. coli* were significantly lower in Little River WWTP sewage than in wastewater from the Johns Creek, Big Creek, and RM Clayton WWTPs (all p<0.05 by post-hoc Tukey Test). Concentrations of 3GC-R *E. coli* were also lower in wastewater from the Utoy Creek WWTP compared to RM Clayton (p<0.05). Concentrations of FQ-R Enterobacterales were significantly lower in Little River WWTP sewage than in wastewater from the Johns Creek and RM Clayton WWTPs (p<0.05).

**Figure 2.**
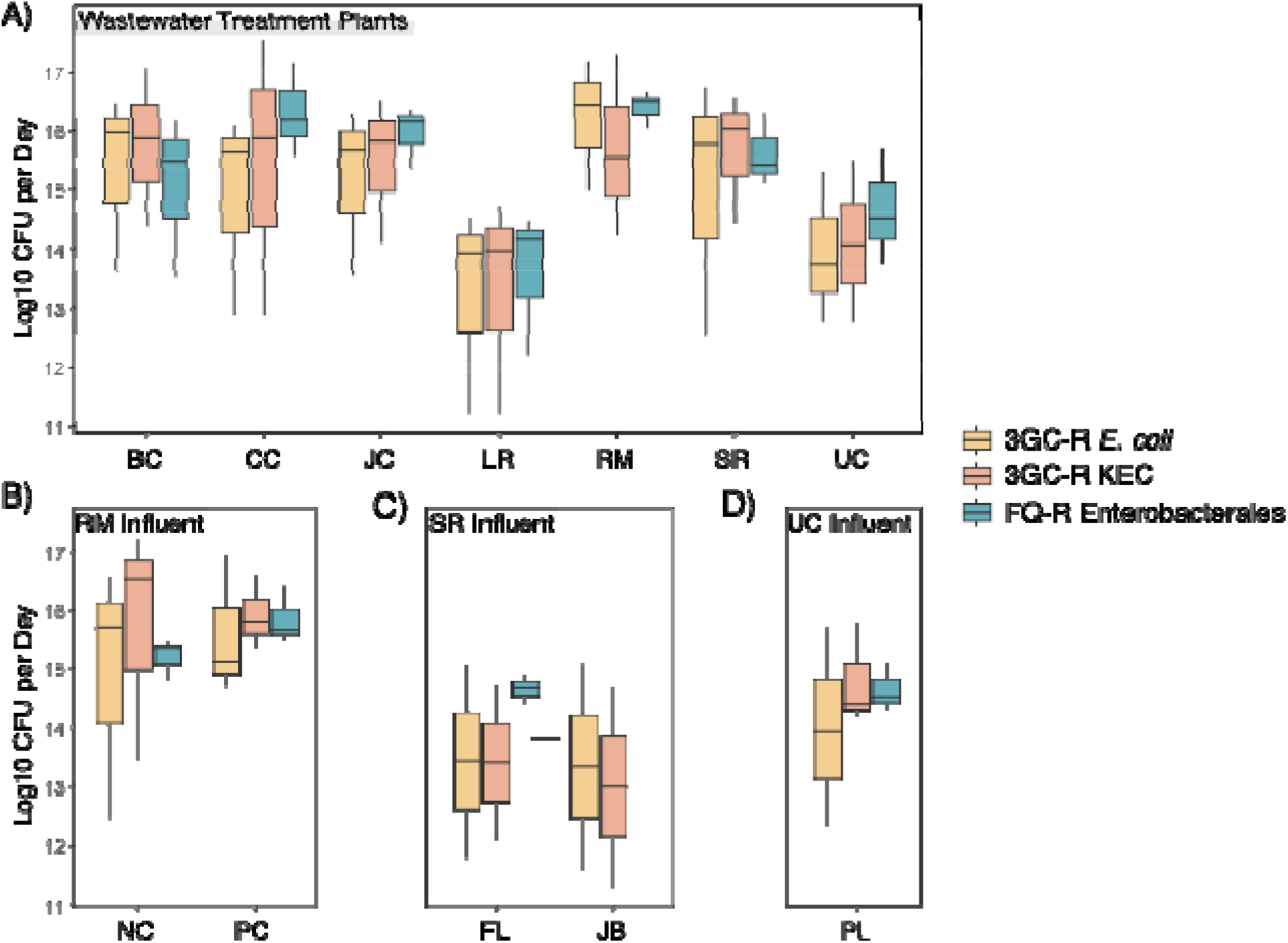
Concentrations of third-generation cephalosporin- and FQ-R Enterobacterales in raw wastewater sampled in December 2022, March 2023, and May 2023 from A) 7 municipal wastewater treatment plants (WWTPs), B) Two influent lines of the RM Clayton WWTP, C) Two influent lines of the South River WWTP, and D) One influent line of the Utoy Creek WWTP. Box plot whiskers depict the full range of concentrations observed, the bottom of each box depicts the 25th percentile, the horizontal line within each box depicts the median, and the top of each box depicts the 75th percentile. Concentrations for WWTPs have been normalized by average daily flow rates. Concentrations for influent lines have been normalized by average monthly flow rates. *<u>Note</u>*: 3GC-R=third-generation cephalosporin-resistant and FQ-R=fluoroquinolone-resistant. BC=Big Creek, CC=Camp Creek, JC=Johns Creek, LR=Little River, RM=RMC Clayton, SR=South River, UC=Utoy Creek, NC=Nancy Creek, PC=Peachtree Creek, FL=Flint, JB=Jonesboro, and PL=Phillip Lee.

Average flow-normalized concentrations of 3GC-R *E. coli* and 3GC-R KEC increased between December and March sampling rounds (p<0.0001 for 3GC-R *E. coli* and p=0.007 for 3GC-R KEC by Tukey test) but not between March and May (p=0.54 for 3GC-R *E. coli* and p=0.99 for 3GC-R KEC by Tukey test), potentially reflecting seasonal differences between winter and spring (**Figure S1**). There was no significant difference in average flow-normalized concentrations of FQ-R Enterobacterales across rounds after accounting for sewershed effects (p=0.07). We observed greater variability in concentrations of 3GC-R *E. coli* and KEC relative to FQ-R Enterobacterales, potentially due to technical challenges in accurately counting pink and blue colonies given substantial overgrowth of other colony morphologies, as others have noted;^20^ or, this could reflect actual variability in wastewater across sampling points.

We detected *bla*_CTX-M-15_ and *bla*_KPC_ in all samples with 16s rRNA normalized concentrations ranging from 4.6 x 10-5 to 0.1 copies/L and 7.1 x 10-4 to 0.5 copies/L, respectively (**Table 2**). Flow-normalized concentrations of *bla*_CTX-M-15_ and *bla*_KPC_ significantly differed across WWTP sewersheds (p<0.05), when accounting for sampling round (**Figure 3**). Again, significantly lower concentrations of both ARGs in Little River WWTP wastewater relative to other WWTPs (i.e., Big Creek, Camp Creek, Johns Creek, South River, and Utoy Creek for *bla*_CTX-M-15_; Camp Creek, Johns Creek, South River, and Utoy Creek for *bla*_KPC_) largely drove these global results, although pairwise differences between other WWTPs were noted.

**Figure 3.**
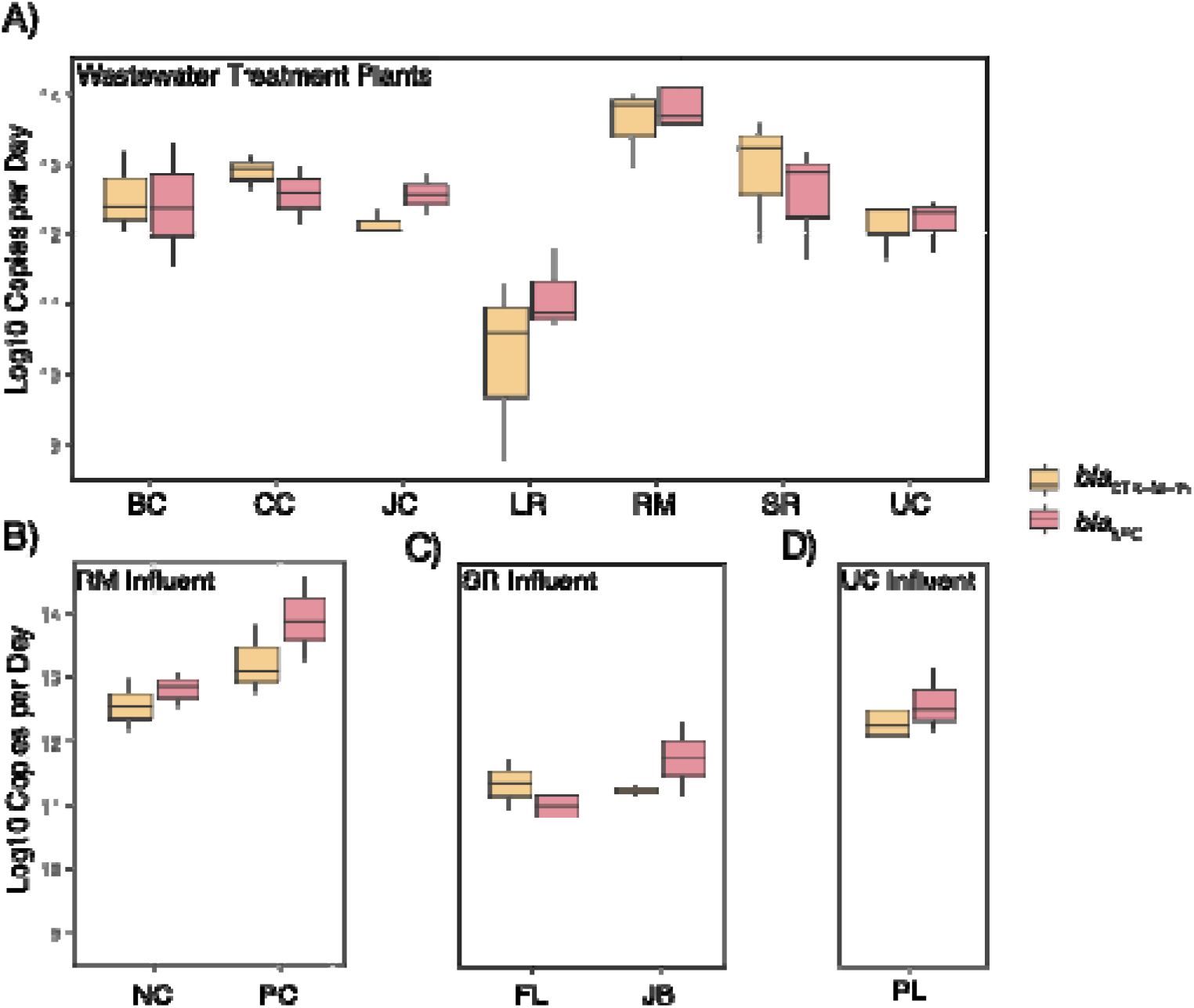
Concentrations of the *bla*_CTX-M-15_ and *bla*_KPC_ genes in raw wastewater sampled in December 2022, March 2023, and May 2023 from A) 7 municipal wastewater treatment plants (WWTPs), B) Two influent lines of the RM Clayton WWTP, C) Two influent lines of the South River WWTP, and D) One influent line of the Utoy Creek WWTP. Box plot whiskers depict the full range of concentrations observed, the bottom of each box depicts the 25th percentile, the horizontal line within each box depicts the median, and the top of each box depicts the 75th percentile. *<u>Note</u>*: Concentrations for WWTPs have been normalized by average daily flow rates. Concentrations for influent lines have been normalized by average monthly flow rates.

**Table 2.**
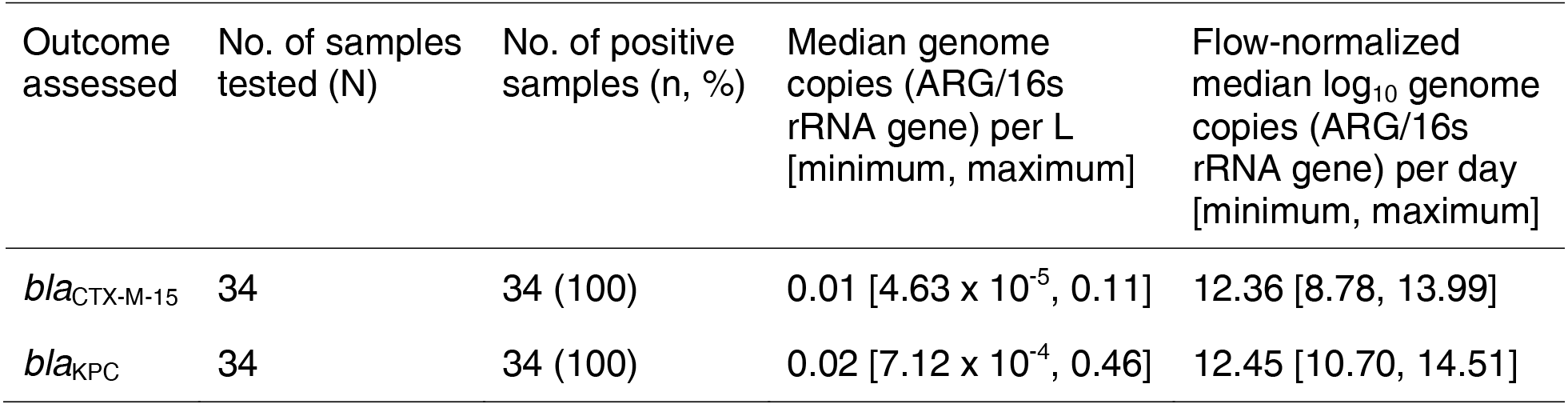
Detection of *bla*_CTX-M-15_ and *bla*_KPC_ in Atlanta area wastewater.

| Outcome assessed | No. of samples tested (N) | No. of positive samples (n, %) | Median genome copies (ARG/16s rRNA gene) per L [minimum, maximum] | Flow-normalized median log<sub>10</sub> genome copies (ARG/16s rRNA gene) per day [minimum, maximum] |
| --- | --- | --- | --- | --- |
| *bla*<sub>CTX-M-15</sub> | 34 | 34 (100) | 0.01 [4.63 x 10<sup>-5</sup>, 0.11] | 12.36 [8.78, 13.99] |
| *bla*<sub>KPC</sub> | 34 | 34 (100) | 0.02 [7.12 x 10<sup>-4</sup>, 0.46] | 12.45 [10.70, 14.51] |

Concentrations of *bla*_CTX-M-15_ were higher in March than in December (p=0.04) but there was no statistical difference in concentrations between March and May (p=0.48); mirroring patterns observed for 3GC-R *E. coli* and 3GC-R KEC. In contrast, concentrations of *bla*_KPC_ did not differ between December and March (p=0.83) but were higher in May than in March (p=0.009) (**Figure S1**).

### Correlations between AR Enterobacterales and ARGs were not consistent

We examined whether *bla*_CTX-M-15_ was associated with concentrations of 3GC-R *E. coli* or KEC to gain insight into the interpretation of *bla*_CTX-M-15_ measurements in wastewater. For both outcomes, the association with *bla*_CTX-M-15_ was found to be homogeneous across sampling rounds; that is, a repeated measures regression model assuming parallelism was selected. thus, we applied linear mixed effects models assuming parallelism and using a diagonal error covariance matrix to evaluate each association. **Table 3** displays the key results of the final selected models, including which error covariance structure was selected via AIC. Although neither relationship was statistically significant at the 0.05 level, we identified trends towards a positive association between *bla*_CTX-M-15_ and both 3GC-R *E. coli* (Estimate=0.39, SE=0.21, Pr > |t| = 0.08) and 3GC-R KEC (Estimate=0.45, SE=0.25, Pr > |t| = 0.09).

**Table 3.**
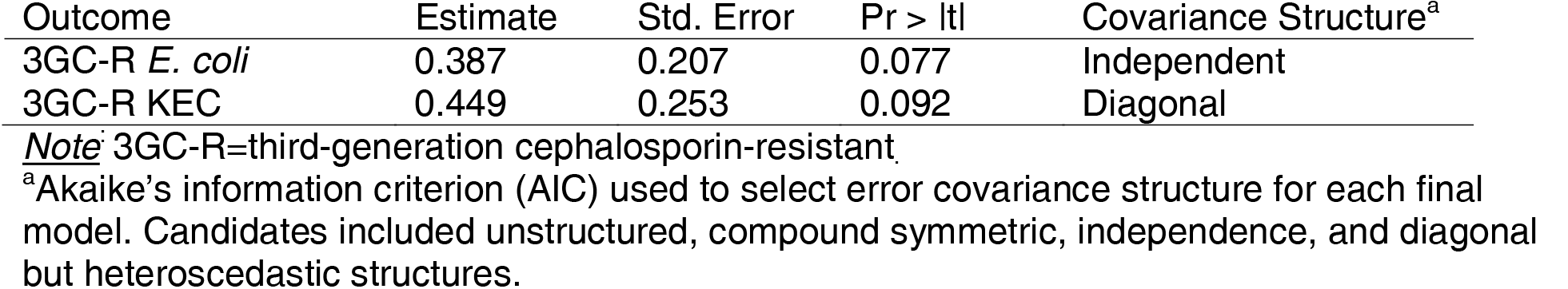
Associations between concentrations of third-generation cephalosporin-resistant (3GCR-R) *E. coli* and 3GC-R KEC in wastewater versus concentrations of the *bla*_CTX-M-15_ gene.

### Distributions of *bla*_CTX-M_ alleles among *3GC-R E. coli* and *K. pneumoniae* were similar across

### sewersheds

More than 70% of tested pink colonies from CHROMagar ESBL were confirmed to be *E. coli*, and approximately 25% of tested blue colonies from CHROMagar KEC were confirmed to be *K. pneumoniae* (**Table S1**). We successfully sequenced the *bla*_CTX-M_ amplicon for 41 3GC-R *E. coli* and 24 3GC-R *K. pneumoniae*. The vast majority of *E. coli* and all *K. pneumoniae* harbored *bla*_CTX-M_-group 1 alleles, although *bla*_CTX-M_-group 9 alleles were also detected among *E. coli* (**Table S7**, **Figure S2**). There were no differences in the proportion of sequenced *E. coli* isolates encoding *bla*_CTX-M_-group 9 across sewersheds (p=0.22 using a 12-sample test for equality of proportions without continuity of correction).

### Sociodemographic characteristics were associated with the detection of AR Enterobacterales in

### wastewater

Sociodemographic characteristics varied widely across sewersheds and among the block groups that comprised them (**Table 4**). We noted sizeable differences across sewersheds in median household income (min=$38,776, max=$175,858), proportion of residents lacking health insurance (min=4.6%, max=16.9%), and racial and ethnic makeup (*e.g.*, 89.5 % non-Hispanic Black in Utoy Creek sewershed versus 6.3% in Little River sewershed).

**Table 4.** Associations between concentrations of third generation cephalosporin-resistant (3GC-R) *E. coli*, 3GC-R KEC, and FQ-R (FQ-R) Enterobacterales in Atlanta wastewater and sewershed-level sociodemographic characteristics, 2022-2023.

| Sewershed-level characteristic | Outcome (log10 CFU/day) | Estimate | Std. Error | Wald test statistic | Pr (>\|W\|) |
| --- | --- | --- | --- | --- | --- |
| % living in crowded households<sup>a</sup> | 3GC-R *E. coli* | 0.48 | 0.11 | 19.10 | 1.24E-05 |
|  | 3GC-R KEC | 0.56 | 0.17 | 10.68 | 1.08E-03 |
|  | FQ-R Enterobacterales | 0.68 | 0.11 | 38.30 | 6.07E-10 |
| % Hispanic<sup>b</sup> | 3GC-R *E. coli* | 0.10 | 0.04 | 8.05 | 4.54E-03 |
|  | 3GC-R KEC | 0.15 | 0.04 | 11.01 | 9.04E-04 |
| % Non-Hispanic Asian<sup>c</sup> | 3GC-R *E. coli* | 0.06 | 0.02 | 14.46 | 1.43E-04 |
|  | 3GC-R KEC | 0.07 | 0.02 | 8.68 | 3.22E-03 |
| % Speaks language other than English at home<sup>d</sup> | 3GC-R *E. coli* | 0.05 | 0.01 | 23.68 | 1.14E-06 |
|  | 3GC-R KEC | 0.05 | 0.01 | 14.60 | 1.33E-04 |

**Note:** Models accounted for repeated measures within each sewershed, sampling round, and number of hospital or long-term care facility beds within each sewershed.
<sup>a</sup>Defined as greater than 1.01 persons/room, not including kitchens or bathrooms. Tabulated as number of owner-occupied households with greater than 1.01 occupants/room divided by total households.
<sup>b</sup>Tabulated as number of persons self-identifying as Hispanic or Latino divided by the total population.
<sup>c</sup>Tabulated as number of persons self-identifying as non-Hispanic Asian alone divided by the total population.
<sup>d</sup>Tabulated as the number of persons 5 years and over who speak a language other than English at home divided by the total population. This variable was highly correlated with percent Hispanic or Latino ( $\rho=0.80$ ) and percent non-Hispanic Asian living in a sewershed ( $\rho=0.95$ ).

We identified four sewershed-level characteristics that were associated with flow-normalized AR Enterobacterales concentrations in untreated influent wastewater when accounting for repeated measures from sewersheds, sampling round, and the number of hospital or LTCF beds within each sewershed, including percent of persons living in crowded households, percent identifying as Hispanic or Latino, percent identifying as non-Hispanic Asian only, and percent speaking another language at home (**Table 4**).

Percent of persons living in crowded households was significantly associated with higher concentrations of all three culture-based outcomes of interest we examined, i.e., FQ-R Enterobacterales (B: 0.68, SE: 0.11, p-value<0.001), 3GC-R *E. coli* (B: 0.48, SE: 0.17, p-value=0.001), and 3GC-R KEC (B: 0.56, SE: 0.11, p-value<0.001). Household crowding was not strongly correlated with any of the other sociodemographic characteristics we evaluated per sewershed (ρ < 0.7 for all pairwise comparisons, **Table S8**), although a moderate correlation with percent speaking a language other than English at home was noted (ρ=0.41).

Percent Hispanic or Latino, percent non-Hispanic Asian, and percent speaking a language other than English at home were all associated with higher concentrations of 3GC-R *E. coli* and 3GC-R KEC in a sewershed’s wastewater (**Table 4**), although effect sizes were small. Percent speaking a non-English language at home was strongly correlated with percent Hispanic or Latino (ρ=0.80, **Table S8**) and percent non-Hispanic Asian (ρ=0.95, **Table S8**) at a sewershed-level, and percent Hispanic or Latino and percent non-Hispanic Asian were moderately correlated with each other (ρ=0.6). Sociodemographic characteristics related to income, insurance status, and higher education status were not strongly correlated with any of these variables at the sewershed level (ρ<0.7 for all).

## Discussion

Antibiotic-resistant bacteria of clinical concern are readily detected in metro Atlanta wastewater. We identified differences in the concentrations of 3GC-R*E coli,* 3GC-R KEC, and FQ-R Enterobacterales in influent wastewater from 12 diverse sewersheds across Atlanta. Our analysis suggests that these differences may be at least partly explained by some of the sociodemographic characteristics of these sewersheds, including percent living in crowded households, percent speaking a non-English language at home, percent Hispanic or Latino, and percent non-Hispanic Asian. Additionally, our findings suggest that the *bla*_CTX-M-15_ gene could be an appropriate surrogate for third generation cephalosporin resistance among human fecal pathogens in wastewater, at least in this setting, which supports the use of this gene in wastewater monitoring. Overall, this study indicates that wastewater sampling could provide new insight into community-level patterns of AR bacteria in urban centers in the United States, though additional work is needed to validate how well these data approximate patterns of human colonization and infection with these bacteria.

Relatively few studies have examined community-level differences in antibiotic-resistant bacteria or ARGs in wastewater from a single urban center. One 2009 study in New York City reported concentrations of seven ARGs over one year across the five boroughs (Manhattan, Queens, Brooklyn, the Bronx, and Staten Island).^21^ Although some differences between boroughs were noted by the study authors, they were not further explored. Some European studies have also examined differences in AR bacteria and ARGs in wastewater from distinct neighborhoods. Among three “socio-spatially” distinct districts of one city in western Germany, a 2021 study^22^ reported consistently higher concentrations of ESBL-producing *E coli* in wastewater from the most disadvantaged community over 1 year. Because the authors’ definition of disadvantage was not clearly defined, however, it is unclear whether some of the notable characteristics we identified in this study - for example, household crowding - may have been more common in the community they identified to be at higher risk in their analysis. Two studies in Basel and Copenhagen also examined AR bacteria or ARGs in influent wastewater from distinct neighborhoods, but differences by neighborhoods’ sociodemographic characteristics were either not identified^23^ or investigated.^24^ To our knowledge, this is the first study to report specific sociodemographic characteristics that may be associated with higher concentrations of clinically-relevant AR bacteria in wastewater. If reflective of the human burden of AR bacteria,^25^ these data may be informative for adapting interventions in community settings. Comparisons to human conization and infection data are needed before wastewater data are leveraged to inform public health practice in this setting.

Proposed wastewater AMR surveillance efforts in the US have largely focused on measuring concentrations of ARGs, including bla_CTX-M-15_ and bla_KPC_. However, these alleles can be harbored by a wide diversity of bacteria that thrive in sewer systems and are not of human origin, obfuscating the public health interpretation of these measurements. In this study, although we consistently detected the bla_KPC_ gene in wastewater, we were unable to reliably culture carbapenem-resistant *E. coli* or KEC from matched samples. Other US studies have also noted challenges in isolating these bacteria for species confirmation from wastewater.^26^ The majority of carbapenem-resistant bacteria in wastewater appear to belong to the family *Aeromonadaceae*,^27,28^ which can grow on chromogenic media meant to be selective and differential for Enterobacterales,^28^ thereby severely complicating our ability to reliably detect carbapenem-resistant *E. coli* or KEC by culture. Carbapenem-resistant Enterobacterales infections are relatively rare in metro Atlanta and only a fraction (9-25%) harbor any carbapenemase gene (**Table S2**). Our detection of low concentrations of the *bla*_KPC_ gene in every sample, therefore, was unusual. Overall, this suggests that the *bla*_KPC_ gene is likely commonly harbored by environmental bacteria, and thus may not be an appropriate surrogate for carbapenem-resistant human fecal pathogens in municipal wastewater. We note that this has implications for the US CDC’s impending plans to implement *bla*_KPC_ monitoring in US wastewaters; specifically, the public health interpretations of this target are unclear.

We accounted for the number of hospital beds and LTCF beds in a sewershed when assessing associations between sociodemographic characteristics and concentrations of AR bacteria in wastewater, but, like others,^29,30^ we found their effects to be near negligible. Given the high volume of antibiotics used in hospital and LTCF settings, both are thought to be important point sources for AR bacteria and ARGs to municipal wastewater. While some studies have found that the presence of hospitals in a sewershed is associated with higher concentrations of AR bacteria in municipal WWTP influent,^23^ others have found that any signal from hospitals appears to “wash out” by the time of arrival at a municipal WWTP.^29,30^ Whether or not hospitals and LTCF meaningfully contribute to the concentrations of AR bacteria in municipal WWTP wastewater may depend on per capita outpatient antibiotic consumption in a given community; healthcare inputs may be more likely to have negligible effects in settings where community antibiotic consumption is relatively high.

Although community-level antibiotic consumption is likely an important driver of the differences we observed in AR bacteria concentrations across sewersheds,^22,31^ we were not able to investigate those differences here. Community-level antibiotic consumption data are not publicly available in the US and existing databases (i.e., IQVIA) do not capture non-prescription use, which is not well-characterized but it thought to be higher among specific minority groups, e.g. Hispanic populations.^32^ Other studies suggest non-prescription use is widespread across racial and ethnic groups in the US.^33^ In theory, higher antibiotic use among Hispanic or non-Hispanic Asian communities could drive the associations we observed between higher proportions of these racial/ethnic groups in Atlanta sewersheds and higher concentrations of 3GC-R *E. coli* and KEC in wastewater. Alternatively, immigrants or their families may also frequently travel to their home countries; travel to Asia and Central and South America is associated with high risk of gut colonization with 3GC-R Enterobacterales,^34–36^ which residents could then be shedding into sewage upon their return. Within the context of this study, it was not possible to distinguish whether international travel, antibiotic consumption patterns, or other factors may have been underpinning the positive associations we observed between concentrations of AR Enterobacterales in wastewater and certain sewershed-level characteristics. However, we note that multiple social determinants of health that have been found to be important risk factors for colonization and infection with AR bacteria in patient-based studies - including low income, low educational attainment, and lack of healthcare access^37^ - were not strongly correlated with the percent of persons living in a sewershed who identified as Hispanic, non-Hispanic Asian, or who spoke a non-English language at home. This suggests that, at least in the US, community-based sampling may yield different insights into the intersections between social determinants of health and antibiotic resistance than are gleaned through healthcare-based studies.

Among the statistically significant sociodemographic characteristics we identified, crowding had the largest effect size. Crowding is a well-known risk factor for the spread of infectious diseases like methicillin-resistant *Staphylococcus aureus*, and has been associated with higher risks for colonization and infection with other pathogens.^37^ The proportion of crowded households in Atlanta sewersheds ranged from 0.2-3.5%. Here, we found that a 1% increase in the number of crowded households in a sewershed was associated with 3 to 4.8 times higher concentrations of 3GC-R*E coli,* 3GC-RKEC, or FQ-R Enterobacterales per day in Atlanta wastewater. Interestingly, household crowding was not strongly correlated with any other sewershed level characteristics that we considered, including poverty or income. The U.S. Department of Housing and Urban Development (HUD) defines crowding as >1.01 persons/room, excluding bathrooms and kitchens. Tolerance or preference for living in multigenerational or multifamily housing that might be considered “crowded” by HUD’s definition can vary according to cultural background, and may occur irrespective of a household’s income status. Given that there is no clear evidence that crowding is associated with higher rates of antibiotic consumption, the underlying drivers of the associations we observed merit further study.

Further work is needed to determine whether the sewershed-level differences we observed in wastewater are reflective of population-level patterns of colonization or infection with AR bacteria. This is challenging for several reasons. First, the majority of AR human fecal bacteria excreted in wastewater are likely commensal organisms. However, data from non healthcare-associated populations on the frequency of colonization with AR pathogens is rare in the United States, which limits the ability to perform such comparisons. Second, fecal bacteria like *E. coli* and *K. pneumoniae*, which were of particular interest in this study, may also originate from non-human sources including domestic pets,^38^ wild animals,^39^ and food waste,^40^ although the relative importance of these inputs is unclear in Atlanta. Livestock may also be a major contributor of AR fecal bacteria in other settings, although, no animal farms, slaughtering plants, or meat processing plants are permitted to discharge waste into the sewersheds we sampled. Third, AR bacteria could also replicate in sewer systems or previously susceptible fecal pathogens could acquire ARGs through horizontal transfer, particularly if bioavailable antibiotics, heavy metals, or other compounds exerting selection pressure are cooccurring in the system, although there is little experimental evidence for this.^41^ Despite these challenges, multiple studies have identified clinically-relevant AR strains in wastewater,^32^ while others have demonstrated increases in AR among wastewater-derived fecal pathogens over time that match trends in geographically-matched patient populations.^42–44^ Overall, this suggests that wastewater may provide high-level, “average” information about patterns of antibacterial resistance in sewershed populations, even if exact population estimates of AR burden are challenging to calculate given the reasons outlined above. We note that any insight gleaned by wastewater sampling could be especially valuable in metro Atlanta where one in five residents lack health insurance and therefore are unlikely to be captured in any clinical surveillance.

There were additional limitations to this study. First, we only tested wastewater at three timepoints (across two seasons). Substantial variability in AR Enterobacterales concentrations between winter and spring sampling rounds suggests that additional sampling might have allowed us to measure relationships with sewershed-level sociodemographic characteristics with less uncertainty. A recent Swiss study estimated that sampling at least once per month results in a 95% CI width below ±0.5%.^25^ Second, we did not specifically account for recent rainfall or surface temperature in our statistical analyses, although we did account for sampling round in all statistical analyses in an effort to account for differing meteorological conditions across timepoints. Recent studies have found mixed impacts of recent rainfall and temperature on AR Enterobacterales concentrations in wastewater.^25,45–47^ Third, we were unable to consider physical differences in sewer systems, including piping material, conductivity, or the presence of biofilms, that could influence replication of AR Enterobacterales, their persistence and viability in sewers, or their ability to exchange ARGs.^45^ These features could have theoretically influenced differences in AR Enterobacterales concentrations in wastewater, although given that sewershed population size (approximated in our study by flow rate) is a major predictor of ARG and AR bacteria concentrations in wastewater,^31,48^ inputs from community and healthcare settings likely exert greater influence. Finally, we used selective media to identify presumptive *E. coli* and *K. pneumoniae*, and it was not feasible to conduct species confirmation on every colony we counted. However, among the random subset of FQ-R Enterobacterales, 3GC-R *E. coli*, and 3GC-R KEC colonies that we did screen, we confirmed that the majority were species of interest, increasing confidence in our results.

## Conclusion

Wastewater sampling could be a low-cost strategy for identifying community-level differences in AR Enterobacterales across urban centers in the United States. Understanding how accurately the sewershed-level differences we observed mirrors trends in the human population will require robust comparisons with human colonization and infection data. Given major gaps in antibiotic resistance surveillance in the US, wastewater sampling merits further investigation as a novel tool for improving public health.

## Data Availability

Raw data needed to replicate our main analyses are available through the Open Science Framework (OSF): https://osf.io/8fcj6/. Sewershed-level sociodemographic characteristics and flow-normalized concentrations of AR bacteria and ARGs are depicted at: https://4f37k2-cameron7564.shinyapps.io/testmap/.

## Acknowledgements

We acknowledge the work of the Georgia Emerging Infections Program and staff who collected and maintained the CRE dataset. EIP surveillance was funded through the Centers for Disease Control and Prevention Emerging Infection Program [U50CK000485]. We thank Shayna Matson and Dania Hussain for their assistance in completing laboratory analyses. We thank Christine Moe for facilitating collaborations between our team and the City of Atlanta.

## References

1. U.S. Department of Health and Human Services, Centers for Disease Control and Prevention (CDC). Antibiotic Resistance Threats in the United States, 2019 [Internet]. Atlanta, GA; 2019 p. 148. Available from: https://www.cdc.gov/drugresistance/pdf/threats-report/2019-ar-threats-report-508.pdf

2. McGregor JC, Bearden DT, Townes JM, Sharp SE, Gorman PN, Elman MR, Mori M, Smith DH. Comparison of antibiograms developed for inpatients and primary care outpatients. Diagn Microbiol Infect Dis. 2013 May;76(1):73–79. PMCID: PMC3658613

3. Shimasaki T, Seekatz A, Bassis C, Rhee Y, Yelin RD, Fogg L, Dangana T, Cisneros EC, Weinstein RA, Okamoto K, Lolans K, Schoeny M, Lin MY, Moore NM, Young VB, Hayden MK, Centers for Disease Control and Prevention Epicenters Program. Increased Relative Abundance of Klebsiella pneumoniae Carbapenemase-producing Klebsiella pneumoniae Within the Gut Microbiota Is Associated With Risk of Bloodstream Infection in Long-term Acute Care Hospital Patients. Clin Infect Dis. 2019 May 30;68(12):2053–2059. PMCID: PMC6541703

4. Ruppé E, Lixandru B, Cojocaru R, Büke C, Paramythiotou E, Angebault C, Visseaux C, Djuikoue I, Erdem E, Burduniuc O, El Mniai A, Marcel C, Perrier M, Kesteman T, Clermont O, Denamur E, Armand-Lefèvre L, Andremont A. Relative fecal abundance of extended-spectrum-β-lactamase-producing Escherichia coli strains and their occurrence in urinary tract infections in women. Antimicrob Agents Chemother. 2013 Sep;57(9):4512–4517. PMCID: PMC3754361

5. Valencia D. Notes from the Field: The National Wastewater Surveillance System’s Centers of Excellence Contributions to Public Health Action During the Respiratory Virus Season — Four U.S. Jurisdictions, 2022–23. MMWR Morb Mortal Wkly Rep [Internet]. 2023 [cited 2023 Dec 8];72. Available from: https://www.cdc.gov/mmwr/volumes/72/wr/mm7248a4.htm

6. Shrider E, Creamer J. Poverty in the United States: 2022 [Internet]. U.S. Census Bureau; 2023 Sep. Report No.: P60-280. Available from: https://www.census.gov/content/dam/Census/library/publications/2023/demo/p60-280.pdf

7. Molina F, López-Acedo E, Tabla R, Roa I, Gómez A, Rebollo JE. Improved detection of Escherichia coli and coliform bacteria by multiplex PCR. BMC Biotechnol. 2015 Jun 4;15:48. PMCID: PMC4453288

8. Barbier E, Rodrigues C, Depret G, Passet V, Gal L, Piveteau P, Brisse S. The ZKIR Assay, a Real-Time PCR Method for the Detection of Klebsiella pneumoniae and Closely Related Species in Environmental Samples. Appl Environ Microbiol. 2020 Mar 18;86(7):e02711–19. PMCID: PMC7082575

9. Duffy N, Karlsson M, Reses HE, Campbell D, Daniels J, Stanton RA, Janelle SJ, Schutz K, Bamberg W, Rebolledo PA, Bower C, Blakney R, Jacob JT, Phipps EC, Flores KG, Dumyati G, Kopin H, Tsay R, Kainer MA, Muleta D, Byrd-Warner B, Grass JE, Lutgring JD, Rasheed JK, Elkins CA, Magill SS, See I. Epidemiology of extended-spectrum β-lactamase–producing Enterobacterales in five US sites participating in the Emerging Infections Program, 2017. Infection Control & Hospital Epidemiology. Cambridge University Press; 2022 Nov;43(11):1586–1594.

10. Emerging Infections Program, Healthcare-Associated Infections – Community Interface Surveillance Report, Multi-site Gram-negative Surveillance Initiative (MuGSI), Carbapenem-Resistant Enterobacterales Surveillance, 2021 [Internet]. US Centers for Disease Control and Prevention (CDC); Available from: https://www.cdc.gov/hai/eip/pdf/mugsi/2021-CRE-Report-508.pdf

11. Redgrave LS, Sutton SB, Webber MA, Piddock LJV. Fluoroquinolone resistance: mechanisms, impact on bacteria, and role in evolutionary success. Trends in Microbiology. Elsevier; 2014 Aug 1;22(8):438–445. PMID: 24842194

12. Water and Wastewater Master Plans (2020-2025): March 2022 Summary [Internet]. Dekalb County, Georgia: Dekalb County Department of Watershed Managamenet; p. 1–42. Available from: https://www.dekalbcountyga.gov/sites/default/files/users/user541/DC%20Water%20and%20WW%20MP%202020-2050_Summary_Final_03042022.pdf

13. Wang Y, Liu P, VanTassell J, Hilton SP, Guo L, Sablon O, Wolfe M, Freeman L, Rose W, Holt C, Browning M, Bryan M, Waller L, Teunis PFM, Moe CL. When case reporting becomes untenable: Can sewer networks tell us where COVID-19 transmission occurs? Water Research. 2023 Feb;229:119516.

14. U.S. Census Bureau. American Community Survey (ACS) Five-Year Data, 2016-2021. 2022.

15. Walker K, Herman M, Eberwein K. tidycensus: Load US Census Boundary and Attribute Data as “tidyverse” and ‘sf’-Ready Data Frames [Internet]. 2024 [cited 2024 Apr 10]. Available from: https://cran.r-project.org/web/packages/tidycensus/index.html

16. Logan JR, Xu Z, Stults B. Interpolating U.S. Decennial Census Tract Data from as Early as 1970 to 2010: A Longtitudinal Tract Database. Prof Geogr. 2014 Jul 1;66(3):412–420. PMCID: PMC4134912

17. United States Census Bureau. TIGER/Line Shapefiles, 2020, for Block Groups in the United States [Internet]. 2022 [cited 2023 Sep 28]. Available from: https://www.google.com/url?q=https://www2.census.gov/geo/tiger/TIGER2020/BG/&sa=D&source=docs&ust=1702325969128841&usg=AOvVaw1jmbkVuvfdcG4KuCdwL4A6

18. H. Akaike. A new look at the statistical model identification. IEEE Transactions on Automatic Control. 1974 Dec;19(6):716–723.

19. SAS Institute, Inc. SAS/STAT 9.4 User’s Guide. Cary, NC: SAS Institute, Inc; 2013.

20. Milligan EG, Calarco J, Davis BC, Keenum IM, Liguori K, Pruden A, Harwood VJ. A Systematic Review of Culture-Based Methods for Monitoring Antibiotic-Resistant Acinetobacter, Aeromonas, and Pseudomonas as Environmentally Relevant Pathogens in Wastewater and Surface Water. Curr Envir Health Rpt. 2023 Feb 23;10(2):154–171.

21. Joseph SM, Battaglia T, Maritz JM, Carlton JM, Blaser MJ. Longitudinal Comparison of Bacterial Diversity and Antibiotic Resistance Genes in New York City Sewage. mSystems. American Society for Microbiology; 2019 Aug 6;4(4):e00327–19.

22. Schmiege D, Zacharias N, Sib E, Falkenberg T, Moebus S, Evers M, Kistemann T. Antibiotic resistance in wastewater from socio-spatially different communities. European Journal of Public Health. 2021 Oct 1;31(Supplement_3):ckab165.627.

23. Gómez-Sanz E, Bagutti C, Roth JA, Alt Hug M, García-Martín AB, Maurer Pekerman L, Schindler R, Furger R, Eichenberger L, Steffen I, Egli A, Hübner P, Stadler T, Aguilar-Bultet L, Tschudin-Sutter S. Spatiotemporal dissemination of ESBL-producing Enterobacterales in municipal sewer systems: a prospective, longitudinal study in the city of Basel, Switzerland. Front Microbiol [Internet]. Frontiers; 2023 May 12 [cited 2024 May 9];14. Available from: https://www.frontiersin.org/journals/microbiology/articles/10.3389/fmicb.2023.1174336/full

24. Brinch C, Leekitcharoenphon P, Duarte ASR, Svendsen CA, Jensen JD, Aarestrup FM. Long-Term Temporal Stability of the Resistome in Sewage from Copenhagen. mSystems. 2020 Oct 20;5(5):e00841–20. PMCID: PMC7577296

25. Conforti S, Holschneider A, Sylvestre É, Julian TR. Monitoring ESBL-Escherichia coli in Swiss wastewater between November 2021 and November 2022: insights into population carriage. mSphere. American Society for Microbiology; 2024 Apr 12;0(0):e00760–23.

26. Foxman B, Salzman E, Gesierich C, Gardner S, Ammerman M, Eisenberg M, Wigginton K. Wastewater surveillance of antibiotic resistant bacteria for public health action: Potential and Challenges. medRxiv. 2024 Jan 1;2024.03.31.24305136.

27. Mathys DA, Mollenkopf DF, Feicht SM, Adams RJ, Albers AL, Stuever DM, Grooters SV, Ballash GA, Daniels JB, Wittum TE. Carbapenemase-producing Enterobacteriaceae and Aeromonas spp. present in wastewater treatment plant effluent and nearby surface waters in the US. PLOS ONE. Public Library of Science; 2019 Jun 26;14(6):e0218650.

28. Drk S, Puljko A, Dželalija M, Udiković-Kolić N. Characterization of Third Generation Cephalosporin- and Carbapenem-Resistant Aeromonas Isolates from Municipal and Hospital Wastewater. Antibiotics (Basel). 2023 Mar 3;12(3):513. PMCID: PMC10044312

29. Buelow E, Bayjanov JR, Majoor E, Willems RJ, Bonten MJ, Schmitt H, van Schaik W. Limited influence of hospital wastewater on the microbiome and resistome of wastewater in a community sewerage system. FEMS Microbiology Ecology. 2018 Jul 1;94(7):fiy087.

30. Buelow E, Rico A, Gaschet M, Lourenço J, Kennedy SP, Wiest L, Ploy MC, Dagot C. Hospital discharges in urban sanitation systems: Long-term monitoring of wastewater resistome and microbiota in relationship to their eco-exposome. Water Research X. 2020 May 1;7:100045.

31. Sims N, Kannan A, Holton E, Jagadeesan K, Mageiros L, Standerwick R, Craft T, Barden R, Feil EJ, Kasprzyk-Hordern B. Antimicrobials and antimicrobial resistance genes in a one-year city metabolism longitudinal study using wastewater-based epidemiology. Environ Pollut. 2023 Sep 15;333:122020. PMID: 37336345

32. Landers TF, Ferng Y hui, McLoughlin JW, Barrett AE, Larson E. Antibiotic identification, use, and self-medication for respiratory illnesses among urban Latinos. Journal of the American Academy of Nurse Practitioners. 2010 Sep 3;22(9):488–495. PMCID: PMC3058843

33. Zoorob R, Grigoryan L, Nash S, Trautner BW. Nonprescription Antimicrobial Use in a Primary Care Population in the United States. Antimicrob Agents Chemother. 2016 Sep;60(9):5527–5532.

34. Ruppé E, Andremont A, Armand-Lefèvre L. Digestive tract colonization by multidrug-resistant Enterobacteriaceae in travellers: An update. Travel Med Infect Dis. 2018 Feb;21:28–35. PMID: 29155322

35. Worby CJ, Earl AM, Turbett SE, Becker M, Rao SR, Oliver E, Taylor Walker A, Walters M, Kelly P, Leung DT, Knouse M, Hagmann SHF, Ryan ET, LaRocque RC. Acquisition and Long-term Carriage of Multidrug-Resistant Organisms in US International Travelers. Open Forum Infectious Diseases. 2020 Dec 1;7(12):ofaa543.

36. Worby CJ, Sridhar S, Turbett SE, Becker MV, Kogut L, Sanchez V, Bronson RA, Rao SR, Oliver E, Walker AT, Walters MS, Kelly P, Leung DT, Knouse MC, Hagmann SHF, Harris JB, Ryan ET, Earl AM, LaRocque RC. Gut microbiome perturbation, antibiotic resistance, and Escherichia coli strain dynamics associated with international travel: a metagenomic analysis. The Lancet Microbe [Internet]. Elsevier; 2023 Sep 13 [cited 2023 Sep 29];0(0). Available from: https://www.thelancet.com/journals/lanmic/article/PIIS2666-5247(23)00147-7/fulltext PMID: 37716364

37. Blackmon S, Avendano E, Nirmala N, Chan CW, Morin RA, Balaji S, McNulty L, Alemu Argaw S, Doron S, Nadimpalli ML. Socioeconomic status and the risk for colonization or infection with priority bacterial pathogens: a global evidence map [Internet]. 2024 [cited 2024 Jun 5]. Available from: http://medrxiv.org/lookup/doi/10.1101/2024.04.24.24306293

38. Guardabassi L. Pet animals as reservoirs of antimicrobial-resistant bacteria: Review. Journal of Antimicrobial Chemotherapy. 2004 Jul 1;54(2):321–332.

39. Worsley-Tonks KEL, Gehrt SD, Miller EA, Singer RS, Bender JB, Forester JD, McKenzie SC, Travis DA, Johnson TJ, Craft ME. Comparison of Antimicrobial-Resistant Escherichia coli Isolates from Urban Raccoons and Domestic Dogs. Elkins CA, editor. Appl Environ Microbiol. 2021 Jul 13;87(15):e00484–21.

40. Zurfluh K, Nüesch-Inderbinen M, Morach M, Zihler Berner A, Hächler H, Stephan R. Extended-spectrum-β-lactamase-producing Enterobacteriaceae isolated from vegetables imported from the Dominican Republic, India, Thailand, and Vietnam. Applied and environmental microbiology. 2015 May;81(9):3115–20. PMID: 25724954

41. Fahrenfeld N, Bisceglia KJ. Emerging investigators series: sewer surveillance for monitoring antibiotic use and prevalence of antibiotic resistance: urban sewer epidemiology. Environ Sci: Water Res Technol. The Royal Society of Chemistry; 2016 Sep 15;2(5):788–799.

42. Reinthaler FF, Galler H, Feierl G, Haas D, Leitner E, Mascher F, Melkes A, Posch J, Pertschy B, Winter I, Himmel W, Marth E, Zarfel G. Resistance patterns of Escherichia coli isolated from sewage sludge in comparison with those isolated from human patients in 2000 and 2009. J Water Health. 2013 Mar;11(1):13–20. PMID: 23428545

43. Kwak YK, Colque P, Byfors S, Giske CG, Möllby R, Kühn I. Surveillance of antimicrobial resistance among *Escherichia coli* in wastewater in Stockholm during 1 year: does it reflect the resistance trends in the society? International Journal of Antimicrobial Agents. 2015 Jan 1;45(1):25–32.

44. Pärnänen KMM, Narciso-da-Rocha C, Kneis D, Berendonk TU, Cacace D, Do TT, Elpers C, Fatta-Kassinos D, Henriques I, Jaeger T, Karkman A, Martinez JL, Michael SG, Michael-Kordatou I, O’Sullivan K, Rodriguez-Mozaz S, Schwartz T, Sheng H, Sørum H, Stedtfeld RD, Tiedje JM, Giustina SVD, Walsh F, Vaz-Moreira I, Virta M, Manaia CM. Antibiotic resistance in European wastewater treatment plants mirrors the pattern of clinical antibiotic resistance prevalence. Sci Adv. 2019 Mar;5(3):eaau9124. PMCID: PMC6436925

45. Eramo A, Medina WRM, Fahrenfeld NL. Factors associated with elevated levels of antibiotic resistance genes in sewer sediments and wastewater. Environ Sci: Water Res Technol. The Royal Society of Chemistry; 2020 Jun 3;6(6):1697–1710.

46. McLellan SL, Huse SM, Mueller-Spitz SR, Andreishcheva EN, Sogin ML. Diversity and population structure of sewage-derived microorganisms in wastewater treatment plant influent. Environmental Microbiology. 2010;12(2):378–392.

47. Roguet A, Newton RJ, Eren AM, McLellan SL. Guts of the Urban Ecosystem: Microbial Ecology of Sewer Infrastructure. mSystems. American Society for Microbiology; 2022 Jun 28;7(4):e00118–22.

48. Newton RJ, McLellan SL, Dila DK, Vineis JH, Morrison HG, Eren AM, Sogin ML. Sewage reflects the microbiomes of human populations. mBio. 2015 Feb 24;6(2):e02574. PMCID: PMC4358014

