## Supplemental File for "Using wastewater sampling to investigate community-level differences in antibacterial resistance in a major urban center, USA"

**Appendix**

| **List of Figures** | **Page** |
| --- | --- |
| Figure S1: Flow-normalized concentrations of A) AR Enterobacterales and B) ARGs by sampling round, among 34 wastewater samples from 12 metro Atlanta sewersheds, 2022-2023. | 2 |
| Figure S2: Distributions of *bla*_CTX-M-1_ and *bla*_CTX-M-9_ group alleles among *E. coli* and *K. pneumoniae* recovered from Atlanta municipal wastewater. | 3 |
| Figure S3: Number of staffed hospital beds and certified long term care facility (LTCF) beds per sewershed. | 4 |

| **List of Tables** |  |
| --- | --- |
| Table S1: Proportion of suspected *Escherichia coli* or *Klebsiella*, *Enterobacter*, or *Citrobacter* spp. (KEC) detected in Atlanta wastewater confirmed to be *E. coli* or *K. pneumoniae* by endpoint PCR. | 5 |
| Table S2: Proportion of incident carbapenem-resistant Enterobacterales (CRE) infections harboring a carbapenemase gene in the metro Atlanta area, 2012-2022. | 6 |
| Table S3: Primer and probe sequences used for detection of antibiotic resistance genes in Atlanta wastewater using digital PCR. | 7 |
| Table S4: ACS5 variables used to tabulate sewershed-level sociodemographic characteristics of interest. | 8-10 |
| Table S5: Flow rates for each wastewater treatment plant (WWTP) and influent line sampled in this study. | 11 |
| Table S6: Associations between concentrations of third-generation cephalosporin-resistant (3GC) E. coli, 3GC-R KEC, and fluoroquinolone-resistant (FQ-R) Enterobacterales in Atlanta wastewater and number of staffed hospital beds and the number of certified long term care facility (LTCF) beds in a sewershed, 2022-2023. | 12 |
| Table S7: Distributions of *bla*_CTX-M_ group alleles among a random subset of *Escherichia coli* (n=41) and *Klebsiella pneumoniae* (n=24) detected in influent wastewater from 12 metro Atlanta sewersheds, 2023-2023. | 13 |
| Table S8: Sewershed-level sociodemographic characteristics that were strongly correlated, i.e., correlation co-efficient >0.7. | 14-16 |

**Figures**

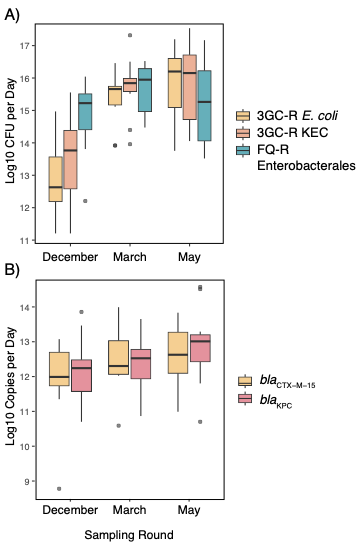

**Figure S1.** Flow-normalized concentrations of **A)** AR Enterobacterales and **B)** ARGs by sampling round, among 34 wastewater samples from 12 metro Atlanta sewersheds, 2022-2023.

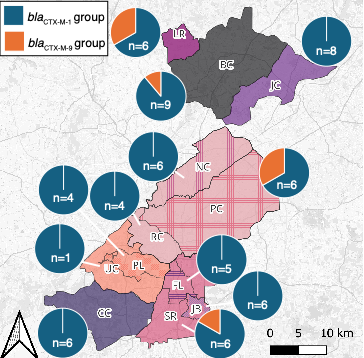

**Figure S2.** Distributions of *bla*_CTX-M-1_ and *bla*_CTX-M-9_ group alleles among *E. coli* and *K. pneumoniae* recovered from Atlanta municipal wastewater.

*Note:* We selected a random subset of up to 3 *bla*_CTX-M_-harboring *E. coli* and 3 *K. pneumoniae* per sewershed, per round, for sequencing. We sequenced the forward strand of the *bla*_CTX-M_ amplicon using Sanger sequencing and determined whether a *bla*_CTX-M_ allele belonging to group 1, 2, 8, 9, 25, 64, 151, or 137 was harbored using BLASTn. We used a 98% identity threshold to identify matches. A total of 65 isolates were sequenced.

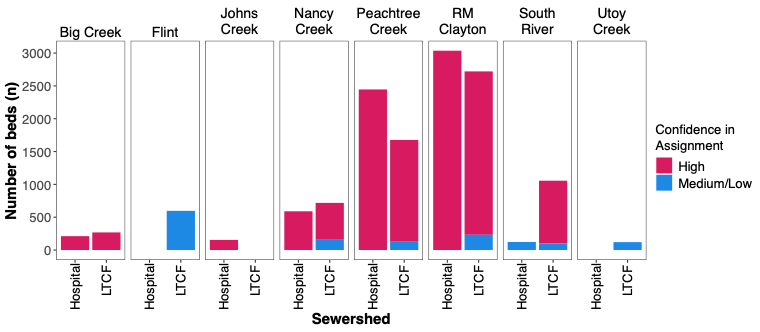

**Figure S3.** Number of staffed hospital beds and certified long term care facility (LTCF) beds per sewershed.

*Note*: High confidence assignments were squarely within the boundaries of a given sewershed; medium or low confidence assignments were relatively close to sewershed boundaries**.** Any sewershed that is not depicted was found to have no hospital nor LTCF beds.

**Tables**

**Table S1**. Proportion of suspected *Escherichia coli* or *Klebsiella*, *Enterobacter*, or *Citrobacter* spp. (KEC) detected in Atlanta wastewater confirmed to be *E. coli* or *K. pneumoniae* by endpoint PCR.

| Culture Media | Colony Phenotype | Number of colonies screened | Confirmed *E. coli*  n (%) | Confirmed *K. pneumoniae*  n (%) |
| --- | --- | --- | --- | --- |
| Chromagar ESBL | Pink (presumptive *E. coli*) | 84 | 61 (73) | -- |
| Chromagar ESBL | Blue (presumptive KEC) | 115 | -- | 27 (23) |
| MacConkey + 4 mg/L ciprofloxacin | Pink or tan (presumptive *E. coli*) | 149 | 103 (69) |  |
| MSuperCarba^a^ | Pink (presumptive *E. coli*) | 5 | 0 | -- |
| MSuperCarba^a^ | Blue (presumptive KEC) | 22 | -- | 1 (5) |

^a^Given the low proportion of presumptive *E. coli* and KEC that were confirmed to be the species of interest, quantitative findings from MSuperCarba are not presented in this manuscript.

*Note*: We were unable to regrow a substantial portion of presumptive carbapenem-resistant *E. coli* or KEC that we had archived on antibiotic-supplemented media for species confirmation, a challenge that other U.S. labs have noted.^1^

**Table S2**. Proportion of incident carbapenem-resistant Enterobacterales (CRE) infections harboring a carbapenemase gene in the metro Atlanta area, 2012-2022.

| Year | 2012 | 2013 | 2014 | 2015 | 2016 | 2017 | 2018 | 2019 | 2020 | 2021 | 2022 |
| --- | --- | --- | --- | --- | --- | --- | --- | --- | --- | --- | --- |
| % CRE harboring a carbapenemase gene | 9.7 | 20.0 | 19.4 | 25.7 | 8.5 | 9.4 | 22.9 | 16.3 | 20.5 | 24.0 | 23.1 |

*Note*: Data provided courtesy of the Georgia Emerging Infections Program.

**Table S3**. Primer and probe sequences used for detection of antibiotic resistance genes in Atlanta wastewater using digital PCR.

| Target gene |  | Sequence (5’ – 3’) | Amplicon size (bp) | Reference |
| --- | --- | --- | --- | --- |
| *bla*_CTX-M-15_ | F    R    Probe | ACCAACGATATCGCGGTGAT    ACATCGCGACGGCTTTCT    6-FAM-TCGTGCGCCGCTG-MGB-NFQ | 101 | Colomer-Lluch et al. (2011a) |
| *bla*_KPC_ | F    R    Probe | GACGGAAAGCTTACAAAAACTGAC    CTTGTCATCCTTGTTAGGCG    FAM-ACTGGGCAGTCGGAGACAAAACCGGA-BHQ1 | 259 | Oliveira et al. (2020)    Poirel et al. (2011)    Oliveira et al. (2020) |
| 16S rRNA | F    R    Probe | GAATGCCACGGTGAATACGTT    TCCCTACGGTTACCTTGTTACG    FAM-CACACCGCCCGTCACACCATGGGAG-BHQ1 | 157 | Oliveira et al. (2020) |

**Table S4.** ACS5 variables used to tabulate sewershed-level sociodemographic characteristics of interest.

| **Variable** | **Numerator** | **Description** | **Denominator** | **Description** |
| --- | --- | --- | --- | --- |
| % Speaks language other than English at home | B99162_003 | Total speak languages other than English for the population 5 years and over | B99162_001 | Total population 5 years and over |
| % Public health insurance | B27010_007 | Total under 19 years of age with Medicaid/means-tested public coverage only | B01003_001 | Total population |
|  | B27010_023 | Total 19-34 years of age with Medicaid/means-tested public coverage only |  |  |
|  | B27010_039 | Total 35-64 years of age with Medicaid/means-tested public coverage only |  |  |
| % No health insurance | B27010_017 | Total under 19 years of age with no health insurance | B01003_001 | Total population |
|  | B27010_033 | Total 19-34 years of age with no health insurance |  |  |
|  | B27010_050 | Total 35-64 years of age with no health insurance |  |  |
|  | B27010_066 | Total 65 years of age and over with no health insurance |  |  |
| Median household income | B19013_001 | Median household income in the past 12 months (2020 inflation-adjusted dollars) |  |  |
| % living in poverty | B29003_002 | Income in the past 12 months below the federal poverty line | B29003_001 | Citizen, Voting-Age population by poverty status |
| % living in crowded households | B25014_005 | Owner Occupied household with 1.01 to 1.50 occupants per room | B25014_001 | Total households by occupants per room |
|  | B25014_006 | Owner Occupied household with 1.51 to 2.00 occupants per room |  |  |
|  | B25014_007 | Owner Occupied household with 2.01 or more occupants per room |  |  |
|  | B25014_011 | Renter Occupied household with 1.01 to 1.50 occupants per room |  |  |
|  | B25014_012 | Renter Owner Occupied household with 1.51 to 2.00 occupants per room |  |  |
|  | B25014_013 | Renter Owner Occupied household with 2.01 or more occupants per room |  |  |
| % Hispanic | B03002_012 | Total Hispanic or Latino | B03002_001 | Total population |
| % Non-Hispanic White | B03002_003 | Total not Hispanic or Latino, White alone | B03002_001 | Total population |
| % Non-Hispanic Black | B03002_004 | Total not Hispanic or Latino, Black alone | B03002_001 | Total population |
| % Non-Hispanic Asian | B03002_006 | Total not Hispanic or Latino, Asian alone | B03002_001 | Total population |
| % completed at least high school^a^ | B15003_003 | Total population 25 years and older with highest level of education attainment being nursery school | B15003_001 | Total population 25 years and older |
|  | B15003_004 | Total population 25 years and older with highest level of education attainment being kindergarten |  |  |
|  | B15003_005 | Total population 25 years and older with highest level of education attainment being 1^st^ grade |  |  |
|  | B15003_006 | Total population 25 years and older with highest level of education attainment being 2^nd^ grade |  |  |
|  | B15003_007 | Total population 25 years and older with highest level of education attainment being 3^rd^ grade |  |  |
|  | B15003_008 | Total population 25 years and older with highest level of education attainment being 4^th^ grade |  |  |
|  | B15003_009 | Total population 25 years and older with highest level of education attainment being 5^th^ grade |  |  |
|  | B15003_010 | Total population 25 years and older with highest level of education attainment being 6^th^ grade |  |  |
|  | B15003_011 | Total population 25 years and older with highest level of education attainment being 7^th^ grade |  |  |
|  | B15003_012 | Total population 25 years and older with highest level of education attainment being 8^th^ grade |  |  |
|  | B15003_013 | Total population 25 years and older with highest level of education attainment being 9^th^ grade |  |  |
|  | B15003_014 | Total population 25 years and older with highest level of education attainment being 10^th^ grade |  |  |
|  | B15003_015 | Total population 25 years and older with highest level of education attainment being 11^th^ grade |  |  |
|  | B15003_016 | Total population 25 years and older with highest level of education attainment being 12^th^ grade, no diploma |  |  |

*Note*: Unless otherwise noted, all variables were tabulated as the sum of all numerator variables divided by the denominator variable.

^a^Tabulated as 1-(Sum of all numerator variables)/denominator variable.

**Table S5**. Flow rates for each wastewater treatment plant (WWTP) and influent line sampled in this study.

|  |  | Flow Rate (MGD) | | |
| --- | --- | --- | --- | --- |
| Location | Type | Dec 2022 | March 2023 | May 2023 |
| South River | Treatment Plant | 19.0 | 35.4 | 23.64 |
| Utoy Creek | Treatment Plant | 28.7 | 38.0 | 30.0 |
| RM Clayton | Treatment Plant | 80.9 | 114.71 | 86.27 |
| Big Creek | Treatment Plant | 21.37 | 26.15 | 16.91 |
| Camp Creek | Treatment Plant | 18.73 | 21.5 | 18.47 |
| Johns Creek | Treatment Plant | 9.78 | 11.6 | 9.02 |
| Little River | Treatment Plant | 2.83 | 1.09 | 0.84 |
| Peachtree Creek | Influent Line | 57.3 | | |
| Nancy Creek | Influent Line | 14.8 | | |
| Flint | Influent Line | 3.2 | | |
| Jonesboro | Influent Line | 1 | | |
| Phillip Lee | Influent Line | 11 | | |

Note: MGD=Million gallons per day. Daily flow rates on the date(s) of sampling were available for WWTPs. Average flow rates across the three months of sampling were available for influent lines. All data were provided by the City of Atlanta and Fulton County.

**Table S6**. Associations between concentrations of third-generation cephalosporin-resistant (3GC) *E. coli*, 3GC-R KEC, and fluoroquinolone-resistant (FQ-R) Enterobacterales in Atlanta wastewater and number of staffed hospital beds and the number of certified long term care facility (LTCF) beds in a sewershed, 2022-2023.

| Sewershed-level characteristic | Outcome  (log10 CFU/day) | Estimate | Std. Error | Wald test statistic | Pr (>\|W\|) |
| --- | --- | --- | --- | --- | --- |
| No. of hospital beds | 3GC-R *E. coli* | 0.0006 | 0.0001 | 42.77 | 6.14E-11 |
|  | 3GC-R KEC | 0.0004 | 0.0001 | 9.99 | 1.57E-03 |
|  | FQ-R Enterobacterales | 0.0004 | 0.0001 | 14.74 | 1.24E-04 |
| No. of LTCF beds | 3GC-R *E. coli* | 0.0007 | 0.0001 | 30.09 | 4.12E-08 |
|  | 3GC-R KEC | 0.0005 | 0.0002 | 6.74 | 9.44E-03 |
|  | FQ-R Enterobacterales | 0.0006 | 0.0002 | 9.66 | 1.88E-03 |

**Table S7.** Distributions of *bla*_CTX-M_ group alleles among a random subset of *Escherichia coli* (n=41) and *Klebsiella pneumoniae* (n=24) detected in influent wastewater from 12 metro Atlanta sewersheds, 2023-2023.

|  | *Escherichia coli* (n=41) | | *Klebsiella pneumoniae* (n=24) | |
| --- | --- | --- | --- | --- |
|  | bla_CTX-M-1_ | bla_CTX-M-9_ | bla_CTX-M-1_ | bla_CTX-M-9_ |
| Big Creek | 5 | 1 | 3 | 0 |
| Camp Creek | 3 | 0 | 3 | 0 |
| Johns Creek | 5 | 0 | 3 | 0 |
| Little River | 3 | 2 | 1 | 0 |
| RM Clayton | 1 | 0 | 3 | 0 |
| South River | 3 | 1 | 1 | 0 |
| Utoy Creek | 0 | 1 |  | 0 |
| Nancy Creek^a^ | 4 | 0 | 2 | 0 |
| Peachtree Creek^a^ | 2 | 2 | 2 | 0 |
| Flint^b^ | 3 | 0 | 2 | 0 |
| Jonesboro^b^ | 3 | 0 | 3 | 0 |
| Phillip Lee^c^ | 2 | 0 | 1 | 0 |

^a^Influent lines of RM Clayton WWTP

^b^Influent lines of SR WWTP

^c^Influent lines of Utoy Creek WWTP

**Table S8**. Sewershed-level sociodemographic characteristics that were strongly correlated, i.e., correlation co-efficient >0.7.

| Variable 1 | Variable 2 | Correlation Coefficient |
| --- | --- | --- |
| At_least_highschool_pct | Median_Household_Income | 0.913 |
| At_least_highschool_pct | non_hispanic_white_pct | 0.85 |
| At_least_highschool_pct | non_hispanic_black_pct | -0.839 |
| At_least_highschool_pct | uninsured_pct | -0.902 |
| At_least_highschool_pct | public_ins_pct | -0.937 |
| At_least_highschool_pct | poverty_pct | -0.939 |
| hispanic_pct | speak_other_lang_pct | 0.803 |
| hispanic_pct | non_hispanic_white_pct | 0.744 |
| hispanic_pct | public_ins_pct | -0.715 |
| hispanic_pct | non_hispanic_black_pct | -0.804 |
| Median_Household_Income | non_hispanic_white_pct | 0.931 |
| Median_Household_Income | At_least_highschool_pct | 0.913 |
| Median_Household_Income | poverty_pct | -0.856 |
| Median_Household_Income | non_hispanic_black_pct | -0.9 |
| Median_Household_Income | uninsured_pct | -0.902 |
| Median_Household_Income | public_ins_pct | -0.913 |
| non_hispanic_asian_pct | speak_other_lang_pct | 0.951 |
| non_hispanic_asian_pct | public_ins_pct | -0.728 |
| non_hispanic_asian_pct | non_hispanic_black_pct | -0.779 |
| non_hispanic_black_pct | public_ins_pct | 0.945 |
| non_hispanic_black_pct | uninsured_pct | 0.878 |
| non_hispanic_black_pct | poverty_pct | 0.826 |
| non_hispanic_black_pct | non_hispanic_asian_pct | -0.779 |
| non_hispanic_black_pct | speak_other_lang_pct | -0.798 |
| non_hispanic_black_pct | hispanic_pct | -0.804 |
| non_hispanic_black_pct | At_least_highschool_pct | -0.839 |
| non_hispanic_black_pct | Median_Household_Income | -0.9 |
| non_hispanic_black_pct | non_hispanic_white_pct | -0.985 |
| non_hispanic_white_pct | Median_Household_Income | 0.931 |
| non_hispanic_white_pct | At_least_highschool_pct | 0.85 |
| non_hispanic_white_pct | hispanic_pct | 0.744 |
| non_hispanic_white_pct | poverty_pct | -0.828 |
| non_hispanic_white_pct | uninsured_pct | -0.895 |
| non_hispanic_white_pct | public_ins_pct | -0.941 |
| non_hispanic_white_pct | non_hispanic_black_pct | -0.985 |
| poverty_pct | uninsured_pct | 0.956 |
| poverty_pct | public_ins_pct | 0.939 |
| poverty_pct | non_hispanic_black_pct | 0.826 |
| poverty_pct | non_hispanic_white_pct | -0.828 |
| poverty_pct | Median_Household_Income | -0.856 |
| poverty_pct | At_least_highschool_pct | -0.939 |
| public_ins_pct | non_hispanic_black_pct | 0.945 |
| public_ins_pct | poverty_pct | 0.939 |
| public_ins_pct | uninsured_pct | 0.926 |
| public_ins_pct | hispanic_pct | -0.715 |
| public_ins_pct | speak_other_lang_pct | -0.726 |
| public_ins_pct | non_hispanic_asian_pct | -0.728 |
| public_ins_pct | Median_Household_Income | -0.913 |
| public_ins_pct | At_least_highschool_pct | -0.937 |
| public_ins_pct | non_hispanic_white_pct | -0.941 |
| speak_other_lang_pct | non_hispanic_asian_pct | 0.951 |
| speak_other_lang_pct | hispanic_pct | 0.803 |
| speak_other_lang_pct | public_ins_pct | -0.726 |
| speak_other_lang_pct | non_hispanic_black_pct | -0.798 |
| uninsured_pct | poverty_pct | 0.956 |
| uninsured_pct | public_ins_pct | 0.926 |
| uninsured_pct | non_hispanic_black_pct | 0.878 |
| uninsured_pct | non_hispanic_white_pct | -0.895 |
| uninsured_pct | At_least_highschool_pct | -0.902 |
| uninsured_pct | Median_Household_Income | -0.902 |
| At_least_highschool_pct | Median_Household_Income | 0.913 |
| At_least_highschool_pct | non_hispanic_white_pct | 0.85 |
| At_least_highschool_pct | non_hispanic_black_pct | -0.839 |
| At_least_highschool_pct | uninsured_pct | -0.902 |
| At_least_highschool_pct | public_ins_pct | -0.937 |
| At_least_highschool_pct | poverty_pct | -0.939 |
| hispanic_pct | speak_other_lang_pct | 0.803 |
| hispanic_pct | non_hispanic_white_pct | 0.744 |
| hispanic_pct | public_ins_pct | -0.715 |
| hispanic_pct | non_hispanic_black_pct | -0.804 |
| Median_Household_Income | non_hispanic_white_pct | 0.931 |
| Median_Household_Income | At_least_highschool_pct | 0.913 |
| Median_Household_Income | poverty_pct | -0.856 |
| Median_Household_Income | non_hispanic_black_pct | -0.9 |
| Median_Household_Income | uninsured_pct | -0.902 |
| Median_Household_Income | public_ins_pct | -0.913 |
| non_hispanic_asian_pct | speak_other_lang_pct | 0.951 |
| non_hispanic_asian_pct | public_ins_pct | -0.728 |

Correlation coefficients generated using the corr() function in R. Variables were created using ACS5 variables described in **Table S4**. Only correlations with correlation co-efficient >0.7 are depicted.
